# Predicting Depressive and Anxiety Symptoms Among Lebanese and Syrian adults in a Suburb of Beirut during the Concurrent Crises: A Population-Based Study

**DOI:** 10.1101/2024.10.25.24316097

**Authors:** Hazar Shamas, Marie-Elizabeth Ragi, Berthe Abi Zeid, Jocelyn DeJong, Stephen J. McCall, with the CAEP Study Group

**Affiliations:** Center for Research on Population and Health, Faculty of Health Sciences, American University of Beirut, Beirut, Lebanon; Department of Health Promotion, Education, and Behavior, University of South Carolina, Columbia, South Carolina, USA; Department of Epidemiology and Population Health, Faculty of Health Sciences, American University of Beirut, Beirut, Lebanon

## Abstract

**Background:** People living in low socioeconomic conditions are more prone to depression and anxiety. This study aimed to develop and internally validate prediction models for depressive and anxiety symptoms in Lebanese adults and Syrian refugees residing in a suburb of Beirut, Lebanon.

**Methods:** This was a population-based study among COVID-19 vulnerable adults in low socioeconomic neighborhoods in Sin-El-Fil, Lebanon. Data were collected through a telephone survey between June and October 2022. The outcomes depressive and anxiety symptoms were investigated for Lebanese and Syrian populations. Depressive and anxiety symptoms were defined as having a PHQ-9 and GAD-7 score of 10 or more respectively. Outcomes’ predictors were identified through LASSO regression, discrimination and model calibrations were assessed using area under curve (AUC) and C-Slope.

**Results:** Of 2,045 participants, 1,322 were Lebanese, 664 were Syrian, and 59 were from other nationalities. Among Lebanese and Syrian populations, 25.3% and 43.9% had depressive symptoms, respectively. Additional predictors for depressive symptoms were not attending school, not feeling safe at all at home, and not having someone to count on in times of difficulty. Not having legal residency documentation for Syrian adults was a context-specific predictor for depressive symptoms. These predictors were similar to that of anxiety symptoms. Both Lebanese and Syrian models had good discriminations and excellent calibrations.

**Conclusion:** This study highlights the main predictors of poor mental health were financial, health, and social indicators for both Lebanese and Syrian adults during the concurrent crisis in Lebanon. Findings emphasise social protection and financial support are required in populations with low socioeconomic status.

**Research in context:** *What is already known on this topic:* The prevalence of depression and anxiety has increased globally. Vulnerable populations, such as refugees and populations of low socioeconomic status, are more prone to depression and anxiety.

*What this study adds:* This study included Lebanese and Syrian adults residing in low socioeconomic status areas of Sin-El-Fil, Lebanon. This is a population-based comparison of the predictors to poor mental health in Lebanon between refugees and Lebanese. The study highlights the need to meet financial, physical, and social needs of individuals to address mental health.

*How this study might affect research, practice, or policy:* The findings of this study highlight the need to reduce financial stress, address physical pain and social isolation, and advocate for Syrian residency documentation to reduce the occurrence of anxiety and depressive symptoms in people living in low socioeconomic conditions.

## Introduction

Mental health disorders, including depressive and anxiety symptoms, were substantial contributors to the Global Burden of Disease (GBD) before the COVID-19 pandemic (1). The World Health Organization (WHO) estimated that the COVID-19 pandemic elevated the prevalence of depression and anxiety by 25% (2). This decline in mental health was multifactorial and attributed to several stressors, including limited social interaction, fear of infection, loss of friends or family, and lack of ‘normal’ routine. The pandemic also resulted in loss of income for populations, particularly those employed in informal sectors, or due to the increased care burden of homeschooling or limited social care for elderly populations (3, 4).

These stressors were disparate between different populations and exacerbated health inequalities with increased prevalence of mental health disorders among populations with low socioeconomic status (SES) (5, 6) and women (7).

Lebanon, a lower-middle income country in the MENA region, is an example of a context with many stressors that may impact well-being. Lebanon has been undergoing a protracted political and economic crisis since 2019, with severe devaluation of the local currency that led to a significant portion of the population moving below the poverty line (8). Additionally, Lebanon hosts the highest number of refugees per capita in the globe, where one quarter of the population are refugees, with an estimated 1.5 million from Syria (9). Unlike refugee populations in other contexts, many refugees in Lebanon live in residential areas alongside the Lebanese host populations (10). Both host populations with low SES and refugee populations have stressors that place them at greater risk of mental health disorders, including lack of employment opportunities, limited social mobility or lack of healthcare access (11). While refugees have the added burden of family member loss from conflict, discrimination, fear of deportation, and psychological traumas (12).

Forced displacement is increasing globally due to conflict and climate-related events (13, 14). Most studies on mental health disorders have explored the risk of mental health disorders either in host or refugee populations separately. In Lebanon, for example, prediction models on the risk of mental health disorders were restricted to older Syrian refugees, widowed Syrian refugee women, and Syrian refugees who experienced ambiguous loss of a close family member (15–17).

In the context of Lebanon, there remains an opportunity to explore the predictors of mental health disorders amongst both host and refugee populations with the aim of identifying the common and disparate stressors of mental disorders to inform service delivery and public health planning. Therefore, the aim of this study is to develop and internally validate prediction models for depressive and anxiety symptoms among Lebanese and Syrian adults residing in a low socioeconomic area, of a suburb in Beirut, Lebanon.

## Methods

### Study design and setting

This study is a nested cross-sectional analysis from the first wave of a longitudinal study. The aim of the parent study was to explore the needs of subpopulations that were vulnerable to the increased risk of COVID-19 infection or its impact in Sin-El-Fil, a suburb east of Beirut, Lebanon (18). Participants in the parent study included: a) adults living in low SES areas b) adults 60 years or older c) Syrian refugees, and d) pregnant women. The study protocol was reviewed and approved by the American University of Beirut Social and Behavioral Sciences Institutional Review Board [Reference: SBS-2021-0268].

### Sampling and study population

The study used a multi-staged stratified sampling design using an area-based sampling (19). The suburb of Sin-E-Fil was stratified into areas of high and low SES, where boundaries were geographically defined based on stakeholder consultation with Sin-El-Fil municipality and non-governmental organisations (NGOs). A household listing exercise was carried out in April 2022 whereby all households in Sin-El-Fil were enumerated face-to-face to complete an eligibility screening survey that determines age, nationality and pregnancy status. This helped identify households and individuals with the specific vulnerability criteria mentioned above (20). Pre-consent was obtained from all households with eligible individuals and phone numbers were collected from consenting households. For the study sample selection, a listing of all eligible household members was generated. A sample of older adults and a sample of adults between 18- 60 years old were randomly selected using proportionate allocation. Due to limited numbers, all Syrian adults and pregnant women were selected. This study population included all participants living in areas of low SES who completed the first wave of data collection conducted between June and October 2022 (n=2,045).

Respondents were contacted to complete a computer assisted telephone survey conducted by trained data collectors where data were entered into SurveyCTO software (Dobility Inc., Cambridge, MA, USA). Verbal informed consent to participate in the oral telephone interview was obtained from selected respondents. Older adults (aged 60 years and older) had their ability to participate in the study assessed using five items altered from the University of California, San Diego, Brief Assessment of Capacity to Consent (UBACC) (21). Participants who scored below 7 where not eligible to participate (Supplemental Table 1).

### Data sources

A questionnaire integrating existing questionnaire modules, community-identified priorities, and adapted to the Sin-El-Fil context was developed in English then translated to Arabic. The survey was created in partnership with representatives of Sin-El-Fil municipality, the Ministry of Public Health (MOPH), and NGOs operating in Sin-El-Fil. The survey was internally piloted to ensure the questions were correctly measuring what the study aimed to measure. Data was monitored weekly with call back checks conducted to ensure accuracy of the collected data where a random set of questions were asked again to 5% of the sample for verification purposes.

### Outcome measures

Depressive and anxiety symptoms were the primary and secondary outcomes of interest in the study respectively. Depressive symptoms were measured through the Patient Health Questionnaire-9 (PHQ-9). PHQ-9 includes 9 items assessing depressive symptoms (Supplemental Table 2). PHQ-9 score ranges from 0 to 27 where each item is scored from 0 (not at all) to 3 (nearly every day) (22). Anxiety symptoms were measured through the General Anxiety Disorder-7 (GAD-7). GAD-7 includes 7 items assessing anxiety symptoms (Supplemental Table 3). The GAD-7 score ranges from 0 to 21 where each item is scored from 0 (not at all) to 3 (nearly every day) (23). The PHQ-9 and GAD-7 had excellent reliability in this population (Cronbach’s alpha=0.89 and 0.95 respectively). A score of 10 or more on the PHQ-9 and GAD-7 indicated the presence of depressive and anxiety symptoms respectively and was used as a cut-off point in this study (24, 25).

### Candidate predictors

Predictors of depressive and anxiety symptoms included in model development were those identified based on depressive and anxiety symptoms literature review (15–17).

For depressive and anxiety symptoms prediction models among Lebanese population, fourteen predictors were included: sex, age, education, marital status, number of chronic illnesses, pain that impact usual activity, household water insecurity, household food insecurity, employment status, having household debt, feeling safe inside home, type of tenure, house eviction notice, number of people to count on in difficult times.

Fifteen predictors were included for the development of depressive and anxiety symptoms prediction models among Syrian population. These predictors were: sex, age, education, marital status, number of chronic illness, pain that impact usual activity, household water insecurity, household food insecurity, employment status, having debt, feeling safe inside home, house eviction notice, number of people to count on in difficult times, cash assistance, and legal residency status characterised by the possession of a valid residency permit. The missing values were missing at random. An indicator variable for each existing variable were generated. A value of 1 was given for every missing observation and a value of 0 was given if observation was not missing. Logit models were then run to test if any of the variables predict whether a given variable is missing (26). Complete case analysis was implemented.

### Statistical analysis

Both outcomes PHQ-9 and GAD-7 were binary (score <10 vs ≥10). Almost all candidate predictors were categorical except for age and number of chronic illnesses which were linearly associated with both outcomes. The analysis accounted for the complex design and non-response. Analysis was weighted according to sampling selection probabilities. Frequency of Lebanese and Syrian participants within each candidate predictor along with their weighted percentages, unadjusted weighted odds ratios (ORs) and 95% confidence intervals (CIs) were calculated. Variables with P-values less than 0.05 were considered statistically significant.

Adaptive Least Absolute Shrinkage and Selection Operator (LASSO) logistic regression model was implemented to identify the predictors of depressive and anxiety symptoms for Lebanese and Syrians (27). All candidate predictors were entered for model development. Separate logistic regressions were run on selected predictors to produce ORs and 95% CIs.

The final models’ performances were estimated through their discrimination abilities using the Area Under the Receiver Operating Curves (AUC) or C-Statistic. A model’s AUC ranges from 0.5 to 1 with an AUC of 1 indicating perfect discrimination. Additionally, the calibration slope (C-Slope) along with their calibration plots were generated to assess model calibration. A model with perfect calibration has a slope of 1, indicating good model performance in future datasets (28). Transparent Reporting of a Multivariable Prediction Model for Individual Prognosis or Diagnosis (TRIPOD) and Strengthening the Reporting of Observational Studies in Epidemiology (STROBE) guidelines were used for reporting (29, 30).

## Results

A total of 2,045 respondents participated in the study of which 1,322 (64.6%) were Lebanese, 664 (32.4%) were Syrian, and 59 (3%) were of other nationalities. Among the Lebanese participants, 25.3% were classified as having depressive symptoms and 30.9% were classified to have anxiety symptoms. The median age for those participants was 50 (IQR:35-63), and 45.5% were males (Table 1 and Supplemental Table 4).

**Table 1.** Characteristics of Lebanese participants and their association with depressive symptoms.

|  | Total |  | PHQ-9<10 |  | PHQ-9≥10 |  | Odds Ratio | (95% CI) |
| --- | --- | --- | --- | --- | --- | --- | --- | --- |
|  | n=1,290 | (%) | n= 966 | (74.7%) | n= 324 | (25.3%) |  |  |
| **Age Median (IQR)** | 50 | (35-63) | 50 | (33-63) | 53 | (39-63) | 1.01 | (1.00-1.01) |
| **Missing** | 8 |  | 8 |  | 0 |  |  |  |
| **Sex** |  |  |  |  |  |  |  |  |
| Male | 584 | (45.5) | 439 | (75.2) | 145 | (24.8) |  |  |
| Female | 706 | (54.5) | 527 | (74.4) | 179 | (25.6) | 1.05 | (0.81-1.35) |
| **Education** |  |  |  |  |  |  |  |  |
| School not attended | 98 | (8.4) | 53 | (54.1) | 45 | (45.9) |  |  |
| School not completed | 574 | (47.6) | 407 | (70.9) | 167 | (29.1) | 0.49 | (0.31-0.75) |
| School completed | 174 | (14.1) | 146 | (83.3) | 28 | (16.7) | 0.24 | (0.13-0.42) |
| Vocational | 115 | (9.3) | 85 | (74.7) | 30 | (25.3) | 0.40 | (0.22-0.72) |
| Higher education | 252 | (20.6) | 208 | (82.9) | 44 | (17.1) | 0.24 | (0.15-0.41) |
| Missing | 77 |  | 67 |  | 10 |  |  |  |
| **Marital status** |  |  |  |  |  |  |  |  |
| Single/ Engaged | 435 | (34.0) | 349 | (80.1) | 86 | (19.9) |  |  |
| Married | 710 | (54.4) | 519 | (73.0) | 191 | (27.0) | 1.49 | (1.11-1.99) |
| Divorced/ Separated/ Widowed | 145 | (11.6) | 98 | (67.4) | 47 | (32.6) | 1.94 | (1.27-2.97) |
| **Pain that impacted usual activity** |  |  |  |  |  |  |  |  |
| No | 926 | (71.5) | 722 | (78.1) | 204 | (21.9) |  |  |
| Yes | 361 | (28.5) | 241 | (66.2) | 120 | (33.8) | 1.81 | (1.38-2.38) |
| Missing | 3 |  | 0 |  | 3 |  |  |  |
| **No. of chronic illness Range (0-6) Median (IQR)** | 0 | (0-1) | 0 | (0-1) | 0 | (0-1) | 1.18 | (1.05-1.32) |
| **Eviction notice** |  |  |  |  |  |  |  |  |
| No | 616 | (47.3) | 451 | (73.0) | 165 | (27.0) |  |  |
| Yes | 79 | (5.9) | 49 | (61.0) | 30 | (39.0) | 1.72 | (1.05-2.83) |
| Owned | 584 | (46.8) | 457 | (78.2) | 127 | (21.8) | 0.75 | (0.58-0.98) |
| Missing | 11 |  | 9 |  | 2 |  |  |  |
| **Household water insecurity** |  |  |  |  |  |  |  |  |
| Secure | 374 | (30.1) | 315 | (84.2) | 59 | (15.8) |  |  |
| Insecure | 876 | (69.9) | 615 | (70.1) | 261 | (29.9) | 2.27 | (1.65-3.12) |
| Missing | 40 |  | 36 |  | 4 |  |  |  |
| **Household food insecurity** |  |  |  |  |  |  |  |  |
| Secure | 419 | (35.0) | 388 | (92.7) | 31 | (7.3) |  |  |
| Insecure | 795 | (65.0) | 529 | (66.2) | 266 | (33.8) | 6.46 | (4.33-9.64) |
| Missing | 76 |  | 49 |  | 27 |  |  |  |
| **Employment status** |  |  |  |  |  |  |  |  |
| No | 713 | (56.2) | 518 | (72.5) | 195 | (27.5) |  |  |
| Yes | 570 | (43.8) | 443 | (77.7) | 127 | (22.3) | 0.76 | (0.58-0.98) |
| Missing | 7 |  | 5 |  | 2 |  |  |  |
| **Having debt** |  |  |  |  |  |  |  |  |
| No | 853 | (73.4) | 693 | (80.8) | 160 | (19.2) |  |  |
| Yes | 321 | (26.6) | 180 | (56.0) | 141 | (44.0) | 3.31 | (2.49-4.39) |
| Missing | 116 |  | 93 |  | 23 |  |  |  |
| **Feeling safe inside home** |  |  |  |  |  |  |  |  |
| Very safe | 729 | (57.3) | 581 | (79.6) | 148 | (20.4) |  |  |
| Somewhat safe | 506 | (39.6) | 355 | (70.2) | 151 | (29.8) | 1.66 | (1.27-2.16) |
| Not safe at all | 40 | (3.1) | 21 | (51.4) | 19 | (48.6) | 3.68 | (1.92-7.05) |
| Missing | 15 |  | 9 |  | 6 |  |  |  |
| **Type of tenure** |  |  |  |  |  |  |  |  |
| Rented | 623 | (48.0) | 440 | (70.2) | 183 | (29.8) |  |  |
| Owned | 633 | (50.9) | 501 | (79.0) | 132 | (21.0) | 0.63 | (0.48-0.81) |
| Other/Hosted | 13 | (1.1) | 11 | (83.8) | 2 | (16.2) | 0.46 | (0.10-2.09) |
| Missing | 21 |  | 14 |  | 7 |  |  |  |
| **Number of people to count on in difficult times** |  |  |  |  |  |  |  |  |
| None | 223 | (17.2) | 144 | (63.4) | 79 | (36.6) |  |  |
| One | 447 | (35.2) | 306 | (68.5) | 141 | (31.5) | 0.80 | (0.57-1.13) |
| Two to five | 580 | (45.0) | 485 | (83.8) | 95 | (16.2) | 0.34 | (0.23-0.48) |
| Six and more | 32 | (2.6) | 24 | (75.5) | 8 | (24.5) | 0.56 | (0.24-1.32) |
| Missing | 8 |  | 7 |  | 1 |  |  |  |

The median age for Syrian participants was 34 (IQR:26-41), of which 50% were males. Of Syrian participants, 43.9% were classified as having depressive symptoms, and 47.2% were classified as having anxiety symptoms (Table 2 and Supplemental Table 5). Interpretations of the unadjusted ORs and 95%CI are presented as supplemental material.

**Table 2.**
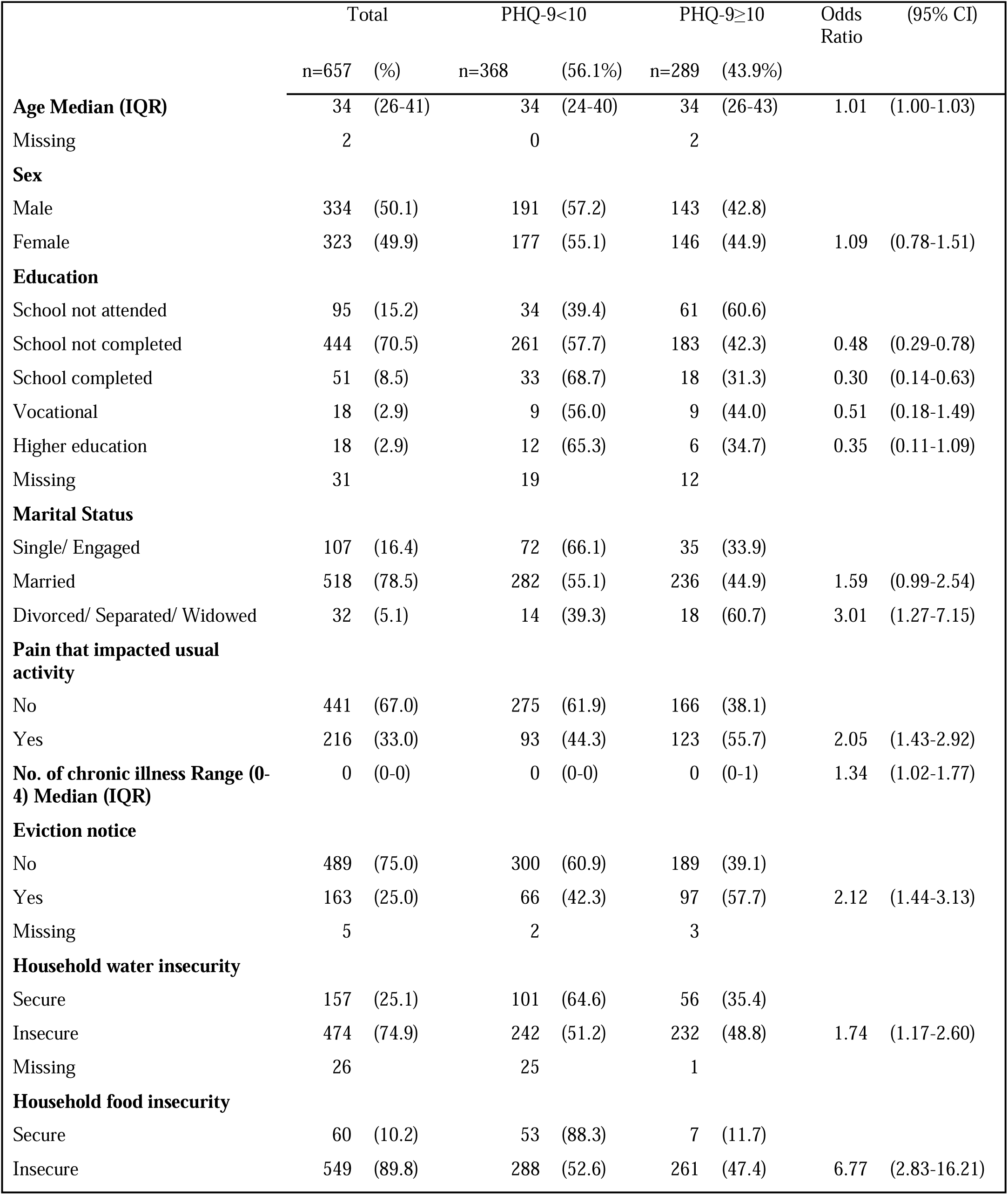
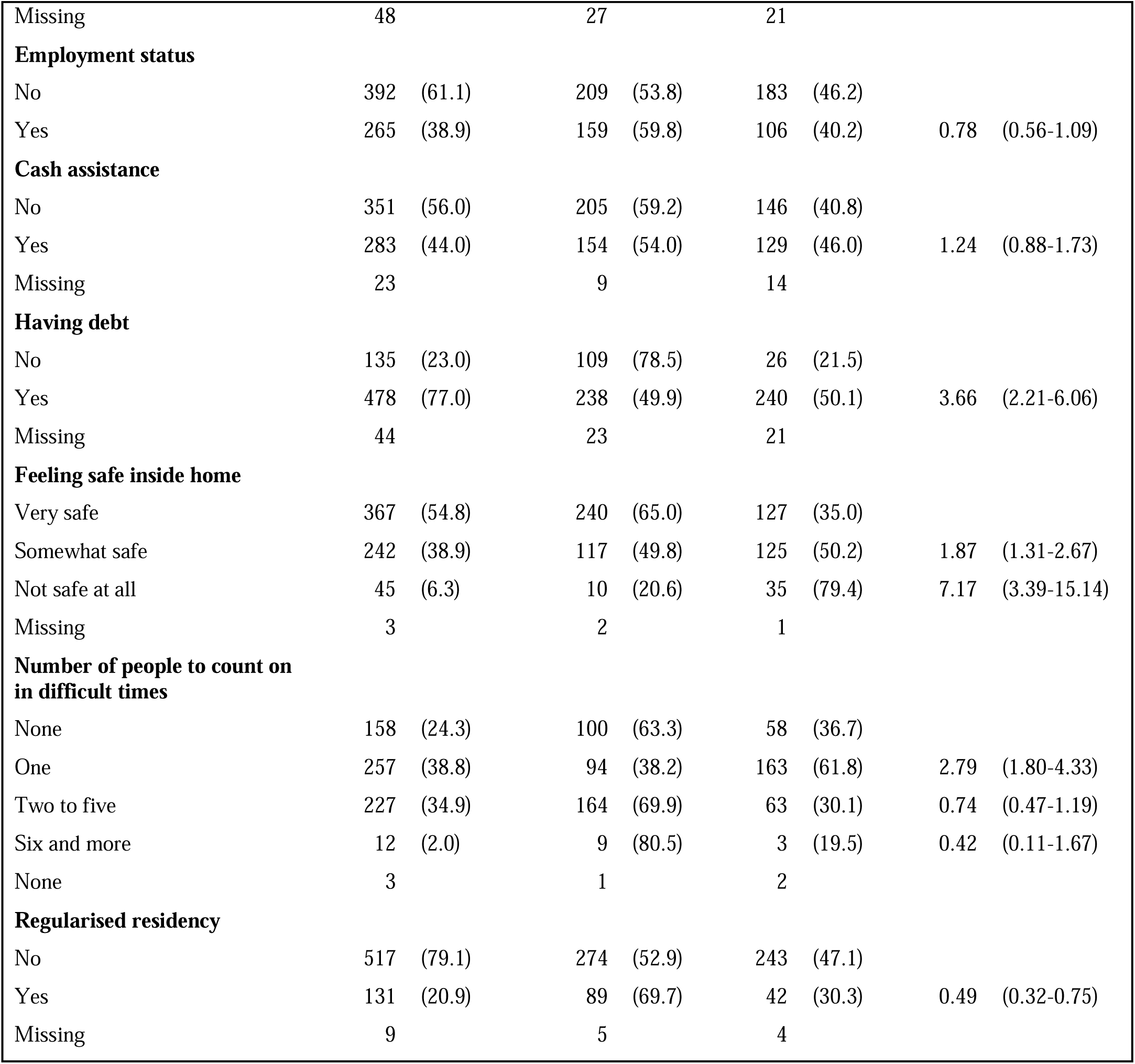
Characteristics of Syrian participants and their association with depressive symptoms.

The Lebanese model predicting depressive symptoms (PHQ-9≥10) retained ten predictors: education, number of chronic illnesses, pain that impacted usual activity, eviction notice, household water insecurity, household food insecurity, having debt, feeling safe inside home, type of housing tenure, and number of people to count on in difficult times. This model had good discrimination (AUC:0.81; 95%CI:0.78-0.84) and excellent calibration (C-Slope:1.06; 95% CI:0.90-1.21) (Figure 1). Predictors of the Lebanese model predicting anxiety symptoms are presented in Supplemental Table 6, model evaluation metrics are presented in results section of the supplemental materials along with Supplemental Figure 1.

**Figure 1.**
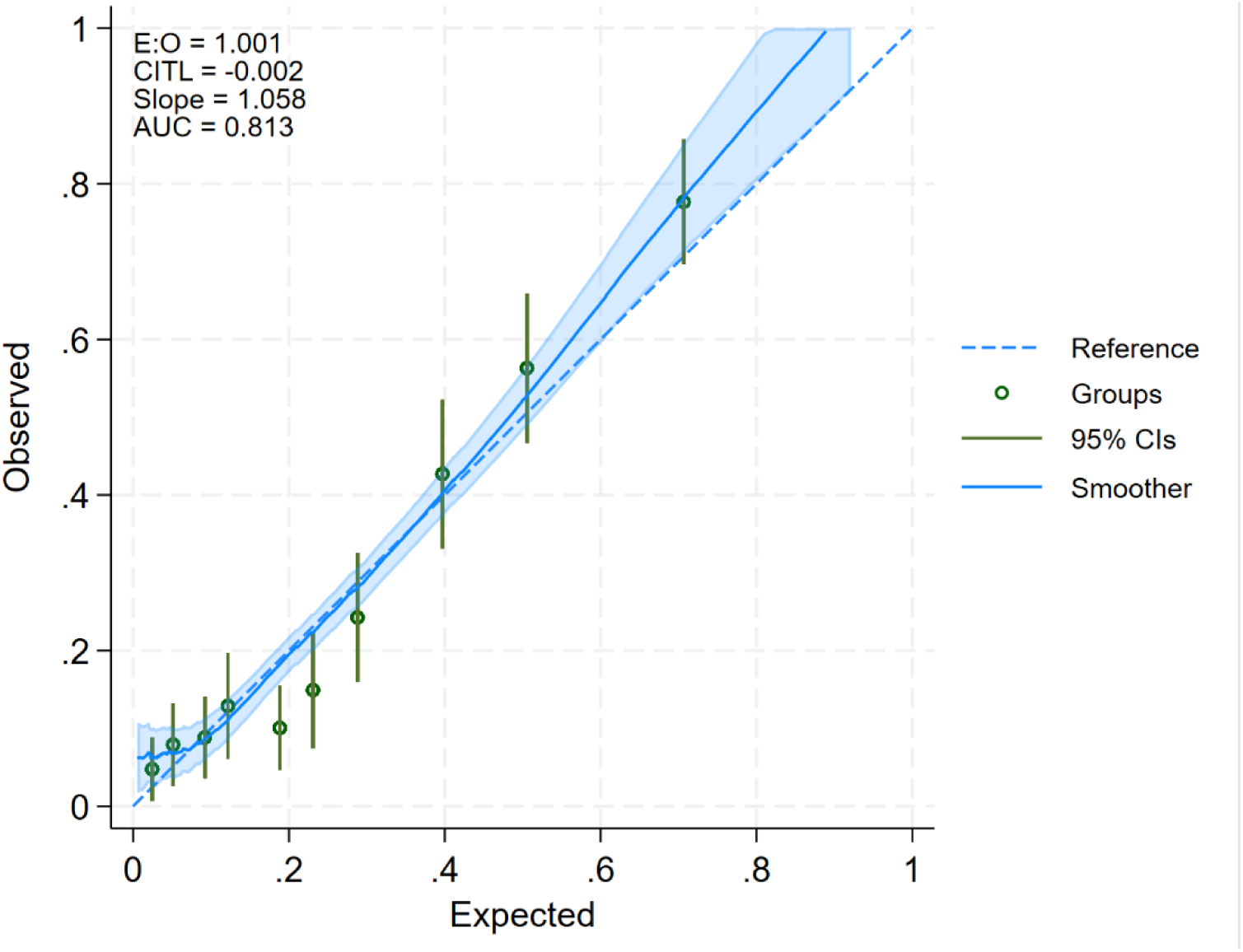
**Calibration plot of Lebanese depressive symptom prediction model**

The Syrian model predicting depressive symptoms retained 10 predictors: education, marital status, number of chronic diseases, pain that impacted usual activity, eviction notice, household food insecurity, having debt, feeling safe inside home, number of people to count on in difficult times, and legal status documentation. This model had good discrimination (AUC:0.83; 95%CI:0.80-0.88) and good calibration slope (C-Slope:1.02; 95%CI:0.84-1.19) (Figure 2).

**Figure 2.**
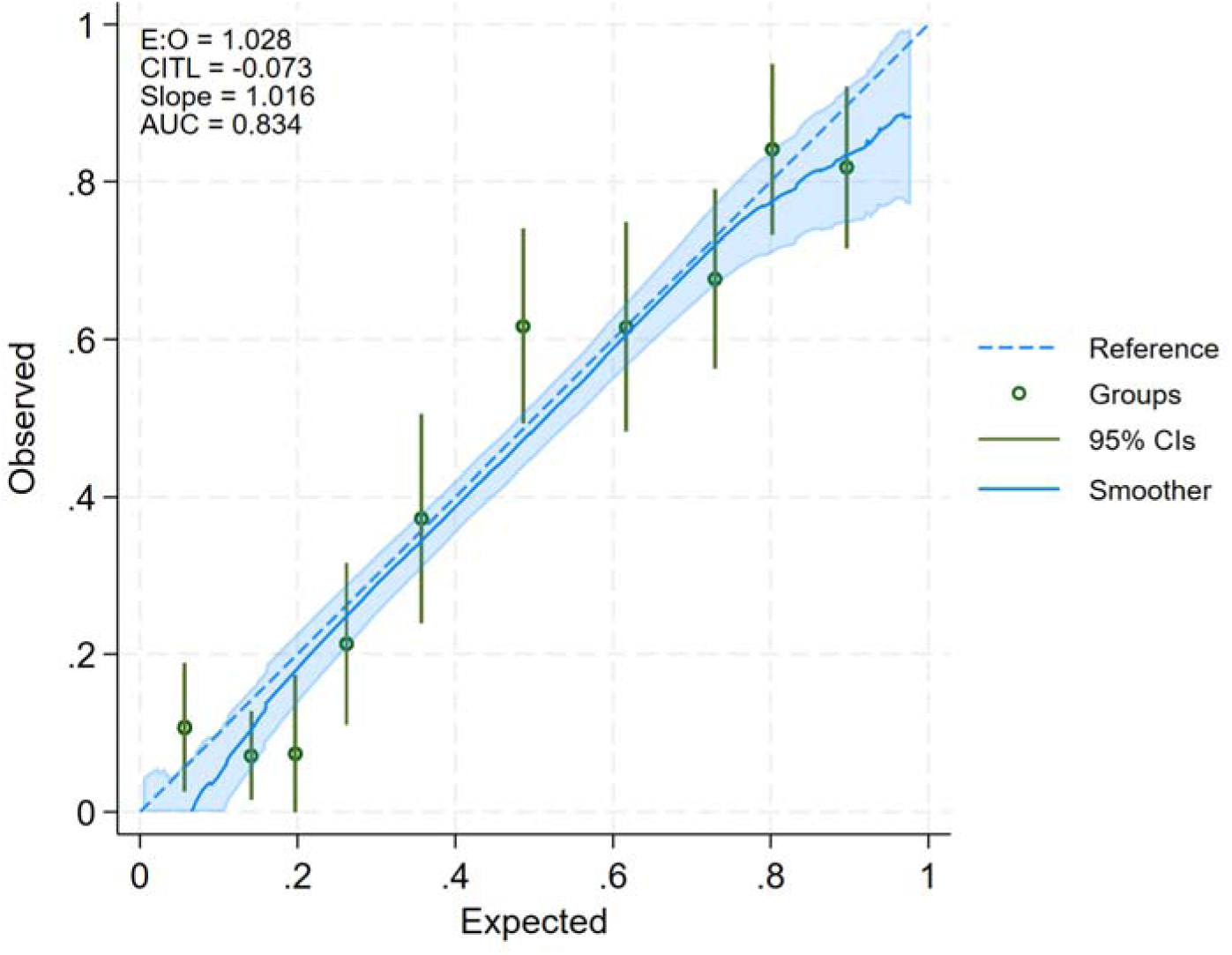
**Calibration plot of Syrian depressive symptom prediction model**

Predictors of the Syrian model predicting anxiety symptoms is presented in Supplemental Table 7, model evaluation metrics are presented in results section of the Supplemental Materials along with Supplemental Figure 2.

Not attending school, having pain that impacted usual activity, having debt, not feeling safe at home, and not having people to count on in difficult times were common predictors of depressive and anxiety symptoms among Lebanese and Syrians. Additional details on Lebanese and Syrian predictors of anxiety symptoms, along with their model evaluation metrics, are presented in the results section of the supplemental.

## Discussion

This study explored predictors of depressive and anxiety symptoms among Lebanese and Syrian populations residing in low socioeconomic areas of Sin-El-Fil, Lebanon. Four prediction models with good discrimination and calibrations were developed. These models aimed to predict depressive and anxiety symptoms among Lebanese and Syrian adults residing in Lebanon. The results of this study also showed high prevalence of depressive and anxiety symptoms in Lebanese and Syrian adults residing in low SES neighborhoods of Sin-El-Fil, Lebanon.

Predictors for a host population from a low socioeconomic area and Syrian refugees in Lebanon were largely similar. The context specific predictor of depressive symptoms for Syrian refugees was lacking a legal residency permit.

Similar to a multicounty representative study, which examined the prevalence of depressive symptoms among refugees and their host population in Kenya, Uganda and Ethiopia, the prevalence of depressive symptoms among refugees was also higher than that of the host population living in the same contexts (31). Moreover, the overall prevalence of depressive symptoms among Syrians in Lebanon was higher than the pooled estimate of depressive symptoms among refugees presented from a global systematic review of 81 studies and 53,458 participants (33.4%; 95%CI: 27.10-39.70) (32).

Similar to other prediction models for poor mental health, this study found that having at least one chronic illness, being in debt, having pain that impacted usual activity, living in a food insecure and water insecure household, and not having a legal residency documentation were selected in the final model (15, 16). Additional predictors included in this study were not attending school, not feeling safe at all at home, and not having someone to count on in times of difficulty.

Similar to other studies, financial predictors of poor mental health included being in debt, food and water insecure, and reception of an eviction notice. Financial stressors contribute to poor mental health through the inability to meet one’s basic needs and overall deteriorating living conditions (33–35). The economic crisis, including the devaluation of currency, loss of savings, and lack of employment opportunities in Lebanon has led many residents to move into poverty (44% of the total population) (36). Many households are unable to meet their basic needs, including their ability to pay rent and obtain enough food which forced many households into debt (37, 38).

Consistent with findings from other studies, being widowed/divorced and having no one to count on through the protracted crisis can intensify loneliness. This compounded isolation often makes it difficult to cope with crisis stressors (39, 40). Thus, with the lack of strong social support networks and prolonged economic crisis, the risk of developing depressive symptoms increases (41, 42).

Aligned with previous national reviews, not feeling safe at home during the COVID-19 pandemic was a predictor of depressive symptoms (43, 44). Staying at home and lockdowns along with the financial stressors elevated the prevalence of psychological abuse and isolation (44). These factors increased the odds of domestic violence among women at home during lockdowns, which independently amplifies the risk of depressive symptoms (43–45).

Similar to other studies, lack of residency documentation was a predictor of mental health disorders (15, 46). Studies have suggested that being undocumented results in migrants experiencing economic uncertainty, being unable to access essential services such as healthcare and being at risk of deportations (46, 47). Therefore, it remains essential to remove the financial and administrative barriers for migrants to obtain and renew their legal documentation.

This study had limitations and strengths. The study sample is representative of individuals living in low SES neighborhoods in Sin-El-Fil, Beirut; however, it may not be generalisable to all Lebanese and Syrian populations residing in Lebanon. On the other hand, this study is one of the only population-based studies on all aged adults in Lebanon. Future research should validate these models on new datasets. Nevertheless, replicable depressive and anxiety symptoms prediction models using innovative machine learning tools with good discrimination and calibration for Lebanese and Syrians residing in Sin-El-Fil were developed.

In conclusion, this study sheds light on the widespread presence of depressive and anxiety symptoms among adults residing in low SES neighborhoods of Sin-El-Fil, Lebanon. It highlights that both refugees and the host population in Lebanon have similar predictors of depression, which are mainly related to socioeconomic characteristics, physical health and social isolation.

The findings of this study provide insight into understanding the risk of mental health disorders, offering a basis for targeting interventions to high-risk populations to reduce the burden in Lebanon.

## Contributors

SJM and HG conceptualised the survey component of study; SJM, HG and MER designed the survey, contribution to study methodology and investigation, and oversaw the data collection. The formal analysis and literature search were conducted by HS. HS, MER, BAZ and SJM drafted the manuscript. SJM supervised MER and HS throughout the project. HS, MER, BAZ, JD, and SJM contributed to the interpretation of the results, edited drafts, and approved the final version of the article. HS, MER and SJM had full access to and verified the raw data. SJM is the guarantor for the study. All authors had access to the study data and had final responsibility for the decision to submit the manuscript for publication.

## Data sharing

The anonymised data can be obtained upon reasonable request from the Center for Research on Population and Health at the American University of Beirut.

## Acknowledgments

This work was funded by the International Development Research Centre (IDRC) – Canada (grant number: 103964; project number: 25941). We would like to thank the “Community Action for Equity in Pandemic preparedness and control” CAEP Study Group for supporting this work. We acknowledge BOT (Bridge. Outsource. Transform) for their assistance in collecting the data required for the success of this study and the study participants for their participation.

## Members of CAEP study group

Aline Germani, Fadi El-Jardali, Hala Ghattas and Nada Melhem

