## Supplemental Material for "Predicting Depressive and Anxiety Symptoms Among Lebanese and Syrian adults in a Suburb of Beirut during the Concurrent Crises: A Population-Based Study"

Affiliations

### METHODS

#### Assessment of capacity to consent

#### Table S1. University of California, San Diego, Brief Assessment of Capacity to Consent (UBACC) items

| 1.What is the purpose of the study that was just described to you? **(2 = the objective of the study)** | 0 | 1 | 2 |
| --- | --- | --- | --- |
| 2.Do you believe this is primarily research or primarily treatment? **(2 = Research)** | 0 | 1 | 2 |
| 3.Do you have to be in this study if you do not want to participate? **(2 = No)** | 0 | 1 | 2 |
| 4.If you participate in this study, what are some of the things that you will be  asked to do? **(2= Answer questions)** | 0 | 1 | 2 |
| 5.Is it possible that being in this study will not have any benefit to you? **(2 = Yes)** | 0 | 1 | 2 |

#### Patient Health Questionnaire-9 (PHQ-9) items

#### Table S2. Patient Health Questionnaire-9 (PHQ-9)

| Over the last 2 weeks, how often have you been bothered by any of the following problems? | Not at all | Several days | More than half of the days | Nearly everyday |
| --- | --- | --- | --- | --- |
| 1.Little interest or pleasure in doing things | 0 | 1 | 2 | 3 |
| 2.Feeling down, depressed, or hopeless | 0 | 1 | 2 | 3 |
| 3.Trouble falling or staying asleep or sleeping too much | 0 | 1 | 2 | 3 |
| 4.Feeling tired or having little energy | 0 | 1 | 2 | 3 |
| 5.Poor appetite or overeating | 0 | 1 | 2 | 3 |
| 6.Feeling bad about yourself – or that you are a failure or have let yourself or your family down | 0 | 1 | 2 | 3 |
| 7.Trouble concentrating on things, such as reading the newspaper or watching television | 0 | 1 | 2 | 3 |
| 8.Moving or speaking so slowly that other people could have noticed? Or the opposite -being so fidgety or restless that you have been moving around a lot more than usual | 0 | 1 | 2 | 3 |
| 9.Thoughts that you would be better off dead or of hurting yourself in some way | 0 | 1 | 2 | 3 |

#### General Anxiety Disorder 7-item scale items

#### Table S3. General Anxiety Disorder 7-item (GAD-7) scale

| Over the last 2 weeks, how often have you been bothered by any of the following problems? | Not at all | Several days | More than half of the days | Nearly everyday |
| --- | --- | --- | --- | --- |
| 1.Feeling nervous, anxious, or on edge | 0 | 1 | 2 | 3 |
| 2.Not being able to stop or control worrying | 0 | 1 | 2 | 3 |
| 3.Worrying too much about different things | 0 | 1 | 2 | 3 |
| 4.Trouble relaxing | 0 | 1 | 2 | 3 |
| 5.Being so restless that it is hard to sit still | 0 | 1 | 2 | 3 |
| 6.Becoming easily annoyed or irritable | 0 | 1 | 2 | 3 |
| 7.Feeling afraid, as if something awful might happen | 0 | 1 | 2 | 3 |

### RESULTS

#### Significant unadjusted ORs representing association between depressive and anxiety symptoms among Lebanese and Syrians along with their 95%CIs

Among the Lebanese participants, factors associated with having depressive symptoms were school not completed compared to school not attended (OR:0.49; 95%CI:0.31-0.75), school completed compared to school not attended (OR:0.24; 95%CI:0.13-0.42), completed vocational school compared to school not attended (OR:0.40; 95%CI:0.22-0.72), completed higher education compared to school not attended (OR:0.24; 95%CI:0.15-0.41), being married compared to being single (OR:1.49; 95%CI: 1.11-1.99), being divorced/ separated/ widowed compared to being single (OR:1.94; 95%CI: 1.27-2.97), having pain that impacted usual activity compared to not having pain that impacted usual activity (OR:1.81; 95%CI:1.38-2.38), number of chronic illnesses (OR:1.18; 95%CI:1.05-1.32), received an eviction notice compared to not receiving an eviction notice (OR:1.72; 95%CI:1.05-2.83), living in an owned home compared to not receiving an eviction notice (OR:0.75; 95%CI:0.58-0.98), household being water insecure compared to being secure (OR:2.27; 95%CI:1.65-3.12), household being food insecure compared to being secure (OR:6.46; 95%CI:4.33-9.64), being employed compared to not being employed (OR:0.76; 95%CI:0.58-0.98), being in debt compared to not having debt (OR:3.31; 95%CI:2.49-4.39), feeling somewhat safe compared to feeling very safe at home (OR:1.66 95%CI:1.27-2.16), not feeling safe at all compared to feeling very safe at home (OR:3.68; 95%CI:1.92-7.05), living in their owned home compared to living in a rented home (OR:0.63; 95%CI:0.48-0.81), and having two to five people as people to count on in difficult times compared to having none (OR:0.34; 95%CI:0.23-0.48) (Table 1).

Factors associated with anxiety symptoms were school not completed compared to school not attended (OR:0.53; 95%CI:0.34-0.81), school completed compared to school not attended (OR:0.35; 95%CI:0.21-0.60), completed vocational school compared to school not attended (OR:0.42; 95%CI:0.24-0.74), completed higher education compared to school not attended (OR:0.27; 95%CI:0.16-0.44), having pain that impacted usual activity compared to not having pain that impacted usual activity (OR:1.52; 95%CI:1.18-1.97), receiving an eviction notice compared to not receiving an eviction notice (OR:1.83; 95%CI:1.13-2.97), household being food insecure compared to being secure (OR:4.07; 95%CI:2.97-5.60), being in debt compared to not having debt (OR:3.97; 95%CI:3.01-5.24), feeling somewhat safe compared to feeling very safe at home (OR:1.96; 95%CI:1.53-2.51), not feeling safe at all compared to feeling very safe at home (OR:3.24; 95%CI:1.69-6.18), living in their owned home compared to living in a rented home (OR:0.75; 95%CI:0.59-0.95), having two to five and six or more people as people to count on in difficult times compared to having none (OR:0.40; 95%CI:0.29-0.56) and (OR:0.37; 95%CI:0.15-0.90) respectively (Supplemental Table 4).

Among the Syrian participants, factors associated with depressive symptoms were school not completed compared to school not attended (OR:0.48; 95%CI:0.29-0.78), school completed compared to school not attended (OR:0.30; 95%CI:0.14-0.63), having pain that impacted usual activity compared to not having pain that impacted usual activity (OR:2.05; 95%CI:1.43-2.92), number of chronic illnesses (OR:1.34; 95%CI:1.02-1.77), had an eviction notice (OR:2.12; 95%CI:1.44-3.13), household being water insecure compared to being secure (OR:1.74; 95%CI:1.17-2.60), household being food insecure compared to being secure (OR:6.77; 95%CI:2.83-16.21), being in debt compared to not having debt (OR:3.66; 95%CI:2.21-6.06), felt somewhat safe (OR:1.87; 95%CI:1.31-267), not feeling safe at all compared to feeling very safe at home (OR:7.17; 95%CI:3.39-15.14), and having regularised residency documents (OR:0.49 95%CI:0.32-0.75) (Table 2).

Factors associated with anxiety symptoms were school completed compared to school not attended (OR:0.44; 95%CI:0.21-0.94), completed higher education compared to school not attended (OR:0.31; 95%CI:0.10-0.92), being married compared to being single (OR:1.87; 95%CI:1.18-2.96), being divorced/ separated/ widowed compared to being single (OR:2.42; 95%CI:1.01-5.81), having pain that impacted usual activity compared to not having pain that impacted usual activity (OR:1.67; 95%CI:1.17-2.37), receiving an eviction notice compared to not receiving an eviction notice (OR:1.66; 95%CI:1.13-2.43), household being water insecure compared to being secure (OR:1.49; 95%CI:1.01-2.20), household being food insecure compared to being secure (OR:3.68; 95%CI:1.90-7.14), being in debt compared to not having debt (OR:3.17; 95%CI:1.99-5.04), feeling somewhat safe compared to feeling very safe at home (OR:1.49; 95%CI:1.05-2.11), not feel safe at all compared to feeling very safe (OR:3.39; 95%CI:1.62-7.06), having one person to count on in difficult times compared to having none (OR:2.25; 95%CI:1.46-3.47), and having six or more people as people to count on in difficult times compared to none (OR:0.20; 95%CI:0.04-0.99) (Supplemental Table 5).

#### Table S4. Characteristics of Lebanese participants and their association with anxiety symptoms

|  | Total |  | GAD-7<10 | | GAD-7≥10 | | Odds Ratio | (95%CI) |
| --- | --- | --- | --- | --- | --- | --- | --- | --- |
|  | n=1,301 | (%) | n=899 | (69.1%) | n=402 | (30.9%) |  |  |
| **Age Median (IQR)** | 50 | (35-63) | 50 | (34-63) | 51 | (36-63) | 1.00 | (1.00-1.01) |
| Missing | 8 |  | 6 |  | 2 |  |  |  |
| **Sex** |  |  |  |  |  |  |  |  |
| Male | 591 | (45.6) | 409 | (69.2) | 182 | (30.8) |  |  |
| Female | 710 | (54.4) | 490 | (69.1) | 220 | (30.9) | 1.00 | (0.79-1.27) |
| **Education** |  |  |  |  |  |  |  |  |
| School not attended | 101 | (8.6) | 52 | (51.9) | 49 | (48.1) |  |  |
| School not completed | 578 | (47.6) | 387 | (67.1) | 191 | (32.9) | 0.53 | (0.34-0.81) |
| School completed | 175 | (14.1) | 131 | (75.2) | 44 | (24.8) | 0.35 | (0.21-0.60) |
| Vocational | 116 | (9.3) | 83 | (72.1) | 33 | (27.9) | 0.42 | (0.24-0.74) |
| Higher education | 252 | (20.4) | 202 | (80.1) | 50 | (19.9) | 0.27 | (0.16-0.44) |
| Missing | 79 |  | 44 |  | 35 |  |  |  |
| **Marital status** |  |  |  |  |  |  |  |  |
| Single/ Engaged | 437 | (33.9) | 316 | (71.9) | 121 | (28.1) |  |  |
| Married | 718 | (54.6) | 491 | (68.5) | 227 | (31.5) | 1.18 | (0.91-1.54) |
| Divorced/ Separated/ Widowed | 146 | (11.6) | 92 | (64.1) | 54 | (35.9) | 1.44 | (0.96-2.14) |
| **Pain that impacted usual activity** |  |  |  |  |  |  |  |  |
| No | 929 | (71.1) | 666 | (71.7) | 263 | (28.3) |  |  |
| Yes | 370 | (28.9) | 231 | (62.5) | 139 | (37.5) | 1.52 | (1.18-1.97) |
| Missing | 2 |  | 2 |  | 0 |  |  |  |
| **No. of chronic illness Range (0-6) Median (IQR)** | 0 | (0-1) | 0 | (0-1) | 0 | (0-1) | 1.09 | (0.98-1.22) |
| **Eviction notice** |  |  |  |  |  |  |  |  |
| No | 623 | (47.5) | 431 | (69.0) | 192 | (31.0) |  |  |
| Yes | 80 | (5.9) | 45 | (54.9) | 35 | (45.1) | 1.83 | (1.13-2.97) |
| Owned | 588 | (46.7) | 416 | (71.0) | 172 | (29.0) | 0.91 | (0.71-1.17) |
| Missing | 10 |  | 7 |  | 3 |  |  |  |
| **Household water insecurity** |  |  |  |  |  |  |  |  |
| Secure | 380 | (30.3) | 269 | (70.8) | 111 | (29.2) |  |  |
| Insecure | 881 | (69.7) | 594 | (67.5) | 287 | (32.5) | 1.17 | (0.90-1.53) |
| Missing | 40 |  | 36 |  | 4 |  |  |  |
| **Household food insecurity** |  |  |  |  |  |  |  |  |
| Secure | 425 | (35.3) | 369 | (86.8) | 56 | (13.2) |  |  |
| Insecure | 799 | (64.7) | 493 | (61.6) | 306 | (38.4) | 4.07 | (2.97-5.60) |
| Missing | 77 |  | 37 |  | 40 |  |  |  |
| **Employment status** |  |  |  |  |  |  |  |  |
| No | 720 | (56.2) | 484 | (67.3) | 236 | (32.7) |  |  |
| Yes | 575 | (43.8) | 413 | (71.9) | 162 | (28.1) | 0.81 | (0.63-1.03) |
| Missing | 6 |  | 2 |  | 4 |  |  |  |
| **Having debt** |  |  |  |  |  |  |  |  |
| No | 858 | (73.4) | 677 | (78.7) | 181 | (21.3) |  |  |
| Yes | 324 | (26.6) | 155 | (48.2) | 169 | (51.8) | 3.97 | (3.01-5.24) |
| Missing | 119 |  | 67 |  | 52 |  |  |  |
| **Feeling safe inside home** |  |  |  |  |  |  |  |  |
| Very safe | 731 | (57.0) | 553 | (75.7) | 178 | (24.3) |  |  |
| Somewhat safe | 514 | (39.9) | 316 | (61.5) | 198 | (38.5) | 1.96 | (1.53-2.51) |
| Not safe at all | 40 | (3.1) | 20 | (49.1) | 20 | (50.9) | 3.24 | (1.69-6.18) |
| Missing | 16 |  | 10 |  | 6 |  |  |  |
| **Type of tenure** |  |  |  |  |  |  |  |  |
| Rented | 629 | (48.1) | 416 | (65.8) | 213 | (34.2) |  |  |
| Owned | 638 | (50.9) | 458 | (72.0) | 180 | (28.0) | 0.75 | (0.59-0.95) |
| Other | 13 | (1.0) | 10 | (77.3) | 3 | (22.7) | 0.57 | (0.15-2.09) |
| Missing | 21 |  | 15 |  | 6 |  |  |  |
| **Number of people to count on in difficult times** |  |  |  |  |  |  |  |  |
| None | 227 | (17.2) | 134 | (58.5) | 93 | (41.5) |  |  |
| One | 451 | (35.3) | 277 | (61.9) | 174 | (38.1) | 0.87 | (0.63-1.21) |
| Two to five | 583 | (44.8) | 455 | (77.9) | 128 | (22.1) | 0.40 | (0.29-0.56) |
| Six and more | 32 | (2.7) | 25 | (79.1) | 7 | (20.9) | 0.37 | (0.15-0.90) |
| None | 8 |  | 8 |  | 0 |  |  |  |

#### Table S5. Characteristics of Syrian participants and their association with anxiety symptoms

|  | Total | | GAD-7<10 | | GAD-7≥10 | | Odds Ratio | | (95%CI) |
| --- | --- | --- | --- | --- | --- | --- | --- | --- | --- |
|  | n=661 | (%) | n=340 | (52.8%) | n=321 | (47.2%) |  |  | |
| **Age Median (IQR)** | 34 | (26-41) | 34 | (25-41) | 34 | (26-42) | 1.00 | (0.99-1.02) | |
| Missing | 2 |  | 0 |  | 2 |  |  |  | |
| **Sex** |  |  |  |  |  |  |  |  | |
| Male | 335 | (49.9) | 174 | (52.8) | 161 | (47.2) |  |  | |
| Female | 326 | (50.1) | 166 | (52.8) | 160 | (47.2) | 1.00 | (0.72-1.39) | |
| **Education** |  |  |  |  |  |  |  |  | |
| School not attended | 95 | (15.1) | 38 | (43.4) | 57 | (56.6) |  |  | |
| School not completed | 448 | (70.7) | 235 | (52.9) | 213 | (47.1) | 0.68 | (0.42-1.11) | |
| School completed | 51 | (8.4) | 31 | (63.4) | 20 | (36.6) | 0.44 | (0.21-0.94) | |
| Vocational | 18 | (2.9) | 8 | (51.2) | 10 | (48.8) | 0.73 | (0.25-2.14) | |
| Higher education | 18 | (2.9) | 12 | (71.5) | 6 | (28.5) | 0.31 | (0.10-0.92) | |
| Missing | 31 |  | 16 |  | 15 |  |  |  | |
| **Marital status** |  |  |  |  |  |  |  |  | |
| Single/ Engaged | 107 | (16.3) | 69 | (65.8) | 38 | (34.2) |  |  | |
| Married | 522 | (78.7) | 258 | (50.7) | 264 | (49.3) | 1.87 | (1.18-2.96) | |
| Divorced/ Separated/ Widowed | 32 | (5.1) | 13 | (44.3) | 19 | (55.7) | 2.42 | (1.01-5.81) | |
| **Pain that impacted usual activity** |  |  |  |  |  |  |  |  | |
| No | 443 | (66.9) | 249 | (57.1) | 194 | (42.9) |  |  | |
| Yes | 218 | (33.1) | 91 | (44.3) | 127 | (55.7) | 1.67 | (1.17-2.37) | |
| **No. of chronic illness Range (0-4) Median (IQR)** | 0 | (0-0) | 0 | (0-0) | 0 | (0-0) | 0.96 | (0.75-1.22) | |
| **Eviction notice** |  |  |  |  |  |  |  |  | |
| No | 492 | (75.0) | 271 | (56.1) | 221 | (43.9) |  |  | |
| Yes | 164 | (25.0) | 67 | (43.5) | 97 | (56.5) | 1.66 | (1.13-2.43) | |
| Missing | 5 |  | 2 |  | 3 |  |  |  | |
| **Household water insecurity** |  |  |  |  |  |  |  |  | |
| Secure | 157 | (25.0) | 90 | (58.7) | 67 | (41.3) |  |  | |
| Insecure | 478 | (75.0) | 226 | (48.8) | 252 | (51.2) | 1.49 | (1.01-2.20) | |
| Missing | 26 |  | 24 |  | 2 |  |  |  | |
| **Household food insecurity** |  |  |  |  |  |  |  |  | |
| Secure | 60 | (10.2) | 46 | (78.8) | 14 | (21.2) |  |  | |
| Insecure | 553 | (89.8) | 272 | (50.2) | 281 | (49.8) | 3.68 | (1.90-7.14) | |
| Missing | 48 |  | 22 |  | 26 |  |  |  | |
| **Employment status** |  |  |  |  |  |  |  |  | |
| No | 396 | (61.3) | 200 | (52.8) | 196 | (47.2) |  |  | |
| Yes | 265 | (38.7) | 140 | (52.9) | 125 | (47.1) | 1.00 | (0.72-1.39) | |
| **Cash assistance** |  |  |  |  |  |  |  |  | |
| No | 355 | (56.3) | 197 | (57.0) | 158 | (43.0) |  |  | |
| Yes | 283 | (43.7) | 135 | (48.7) | 148 | (51.3) | 1.40 | (1.00-1.95) | |
| Missing | 23 |  | 8 |  | 15 |  |  |  | |
| **Having debt** |  |  |  |  |  |  |  |  | |
| No | 135 | (23.0) | 102 | (75.3) | 33 | (24.7) |  |  | |
| Yes | 482 | (77.0) | 228 | (49.1) | 254 | (50.9) | 3.17 | (1.99-5.04) | |
| Missing | 44 |  | 10 |  | 34 |  |  |  | |
| **Feeling safe inside home** |  |  |  |  |  |  |  |  | |
| Very safe | 367 | (54.5) | 212 | (58.4) | 155 | (41.6) |  |  | |
| Somewhat safe | 244 | (38.9) | 114 | (48.6) | 130 | (51.4) | 1.49 | (1.05-2.11) | |
| Not safe at all | 47 | (6.6) | 12 | (29.3) | 35 | (70.7) | 3.39 | (1.62-7.06) | |
| Missing | 3 |  | 2 |  | 1 |  |  |  | |
| **Number of people to count on in difficult times** |  |  |  |  |  |  |  |  | |
| None | 158 | (24.2) | 90 | (57.7) | 68 | (42.3) |  |  | |
| One | 261 | (39.1) | 94 | (37.7) | 167 | (62.3) | 2.25 | (1.46-3.47) | |
| Two to five | 227 | (34.7) | 145 | (64.7) | 82 | (35.3) | 0.75 | (0.48-1.16) | |
| Six and more | 12 | (2.0) | 10 | (87.0) | 2 | (13.0) | 0.20 | (0.04-0.99) | |
| Missing | 3 |  | 1 |  | 2 |  |  |  | |
| **Legal status documentation** |  |  |  |  |  |  |  |  | |
| No | 520 | (79.1) | 263 | (52.1) | 257 | (47.9) |  |  | |
| Yes | 132 | (20.9) | 74 | (58.0) | 58 | (42.0) | 0.79 | (0.52-1.19) | |
| Missing | 9 |  | 3 |  | 6 |  |  |  | |

#### Table S6. Predictors of depressive and anxiety symptoms among Lebanese adults

| **PHQ-9≥10** | | | | **GAD-7≥10** | | | |
| --- | --- | --- | --- | --- | --- | --- | --- |
| **PHQ-9≥10** | **Codes** | **Penalised Coefficients** | **OR (95%CI)** |  | **Codes** | **Penalised Coefficients** | **OR (95%CI)** |
| **Education** |  |  | 1 | **Age** | - | -0.01 | 0.99 (0.97-1.01) |
| School not attended | 0 | 0.46 | 0.62 (0.36-1.08) | **Education** |  |  |  |
| School not completed | 1 | - | 0.39 (0.18-0.81) | School not attended | 0 | 0.62 | 1 |
| School completed | 2 | -0.18 | 0.56 (0.26-1.21) | School not completed | 1 | - | 0.53 (0.30-0.92) |
| Vocational school | 3 | - | 0.51 (0.27-0.98) | School completed | 2 | - | 0.47 (0.22-0.97) |
| Higher education | 4 | - | 0.62 (0.36-1.08) | Vocational school | 3 | - | 0.41 (0.19-0.89) |
| **No. of chronic illness** | - | 0.05 | 1.09 (0.93-1.28) | Higher education | 4 | -0.23 | 0.36 (0.18-0.72) |
| **Pain impacted usual activity** |  |  |  | **Marital status** |  |  |  |
| No | 0 | - | 1 | Single/ Engaged | 0 | - | 1 |
| Yes | 1 | 0.68 | 1.92 (1.31-2.83) | Married | 1 | - | 1.08 (0.72-1.61) |
| **Eviction notice** |  |  |  | Divorced/ Separated/ Widowed | 2 | 0.15 | 1.38 (0.75-2.54) |
| No | 0 | -0.75 | 11 | **Pain impacted usual activity** |  |  |  |
| Yes | 1 | - | 1.79 (0.89-3.60) | No | 0 | - | 1 |
| Owned | 2 | - | 4.39 (1.40-13.7) | Yes | 1 | 0.78 | 2.28 (1.57-3.31) |
| **Household water insecurity** |  |  |  | **Eviction notice** |  |  |  |
| Secure | 0 | - | 1 | No | 0 | -0.65 | 1 |
| Insecure | 1 | 0.56 | 1.92 (1.27-2.90) | Yes | 1 | - | 1.66 (0.82-3.32) |
| **Household food insecurity** |  |  |  | Owned | 2 | - | 3.21 (1.23-7.94) |
| Secure | 0 | - | 1 | **Household water insecurity** |  |  |  |
| Insecure | 1 | 1.81 | 6.18 (3.85-9.93) | Secure | 0 | - | 1 |
| **Having debt** |  |  |  | Insecure | 1 | 0.18 | 1.29 (0.88-1.89) |
| No | 0 | - | 1 | **Household food insecurity** |  |  |  |
| Yes | 1 | 0.91 | 261 (1.83-3.72) | Secure | 0 | - | 1 |
| **Feeling safe inside home** |  |  |  | Insecure | 1 | 1.69 | 5.41 (3.51-8.34) |
| Very safe | 0 | -0.63 | 1 | **Having debt** |  |  |  |
| Somewhat safe | 1 | - | 1.94 (1.37-2.76) | No | 0 | - | 1 |
| Not safe at all | 2 | 0.34 | 3.62 (1.51-8.66) | Yes | 1 | 1.13 | 3.14 (2.21-4.46) |
| **Type of tenure** |  |  |  | **Feeling safe inside home** |  |  |  |
| Rent | 0 | 0.89 | 1 | Very safe | 0 | -0.76 | 1 |
| Owned | 1 | - | 0.20 (0.06-0.62) | Somewhat safe | 1 | - | 2.11 (1.50-2.96) |
| Other/ Hosted | 2 | -0.23 | 0.15 (0.01-1.26) | Not safe at all | 2 | 0.35 | 3.53 (1.49-8.34) |
| **Number of people to count on in difficult times** |  |  |  | **Type of tenure** |  |  |  |
| Zero | 0 | 0.32 | 1 | Rent | 0 | 0.51 | 1 |
| One | 1 | - | 0.68 (0.38-1.00) | Owned | 1 | - | 0.38 (0.15-0.96) |
| Two to five | 2 | -1.05 | 0.22 (0.13-0.35) | Other/ Hosted | 2 | - | 0.49 (0.12-2.08) |
| Six or more | 3 | - | 0.83 (0.29-2.40) | **Number of people to count on in difficult times** |  |  |  |
|  |  |  |  | Zero | 0 | 0.21 | 1 |
|  |  |  |  | One | 1 | - | 0.74 (0.46-1.19) |
|  |  |  |  | Two to five | 2 | - | 0.26 (0.16-0.42) |
|  |  |  |  | Six or more | 3 | -1.03 | 0.61 (0.21-1.74) |

#### Table S7. Predictors of depressive and anxiety symptoms among Syrian adults

| **PHQ-9≥10** | | | | **GAD-7≥10** | | | |
| --- | --- | --- | --- | --- | --- | --- | --- |
|  | **Codes** | **Penalised Coefficients** | **OR (95%CI)** |  | **Codes** | **Penalised Coefficients** | **OR (95%CI)** |
| **Education** |  |  |  | **Education** |  |  |  |
| School not attended | 0 | 0.81 | 1 | School not attended | 0 | - | 1 |
| School not completed | 1 | - | 0.44 (0.23-0.84) | School not completed | 1 | - | 0.73 (0.44-1.23) |
| School completed | 2 | -0.36 | 0.25 (0.09-0.72) | School completed | 2 | -0.32 | 0.42 (0.18-0.96) |
| Vocational school | 3 | - | 0.57 (0.14-1.35) | Vocational school | 3 | - | 1.19 (0.37-3.84) |
| Higher education | 4 | - | 0.69 (0.16-2.90) | Higher education | 4 | - | 0.42 (0.12-1.47) |
| **Marital status** |  |  |  | **Pain impact usual activity** |  |  |  |
| Single/ Engaged | 0 | - | 1 | No | 0 | - | 1 |
| Married | 1 | - | 1.10 (0.57-2.09) | Yes | 1 | 0.39 | 1.55 (1.05-2.27) |
| Divorced/ Separated/ Widowed | 2 | 0.48 | 2.06 (0.62-6.79) | **Having debt** |  |  |  |
| **No. of chronic illness** |  | 0.21 | 1.31 (0.93-1.82) | No | 0 | - | 1 |
| **Pain impacted usual activity** |  |  |  | Yes | 1 | 1.26 | 2.87 (1.75-4.70) |
| No | 0 | - | 1 | **Feeling safe inside home** |  |  |  |
| Yes | 1 | 0.52 | 1.69 (1.05-2.70) | Very safe | 0 | -0.35 | 1 |
| **Eviction notice** |  |  |  | Somewhat safe | 1 | - | 1.29 (0.87-1.91) |
| No | 0 | - | 1 | Not safe at all | 2 | 0.91 | 3.65 (1.67-7.94) |
| Yes | 1 | 0.45 | 1.61 (0.97-2.68) | **Number of people to count on in difficult times** |  |  |  |
| **Household food insecurity** |  |  |  | Zero | 0 | - | 1 |
| Secure | 0 | - | 1 | One | 1 | 0.50 | 2.70 (1.66-4.37) |
| Insecure | 1 | 1.61 | 5.19 (1.82-14.80) | Two to five | 2 | -0.65 | 0.73 (0.44-1.23) |
| **Having debt** |  |  |  | Six or more | 3 | - | 0.40 (0.07-2.13) |
| No | 0 | - | 1 |  |  |  |  |
| Yes | 1 | 1.47 | 4.09 (2.12-7.87) |  |  |  |  |
| **Feeling safe inside home** |  |  |  |  |  |  |  |
| Very safe | 0 | -0.82 | 1 |  |  |  |  |
| Somewhat safe | 1 | - | 2.21 (1.36-3.59) |  |  |  |  |
| Not safe at all | 2 | 0.35 | 3.58 (1.47-8.72) |  |  |  |  |
| **Number of people to count on in difficult times** |  |  |  |  |  |  |  |
| Zero | 0 | - | 1 |  |  |  |  |
| One | 1 | 0.95 | 2.67 (1.46-4.90) |  |  |  |  |
| Two to five | 2 | -1.05 | 0.32 (0.16-0.60) |  |  |  |  |
| Six or more | 3 | - | 0.96 (0.17-5.28) |  |  |  |  |
| **Legal status documentation** |  |  |  |  |  |  |  |
| No | 0 | - | 1 |  |  |  |  |
| Yes | 1 | -0.54 | 0.56 (0.31-1.01) |  |  |  |  |

#### Figure S1. Calibration plot of Lebanese anxiety symptom prediction model

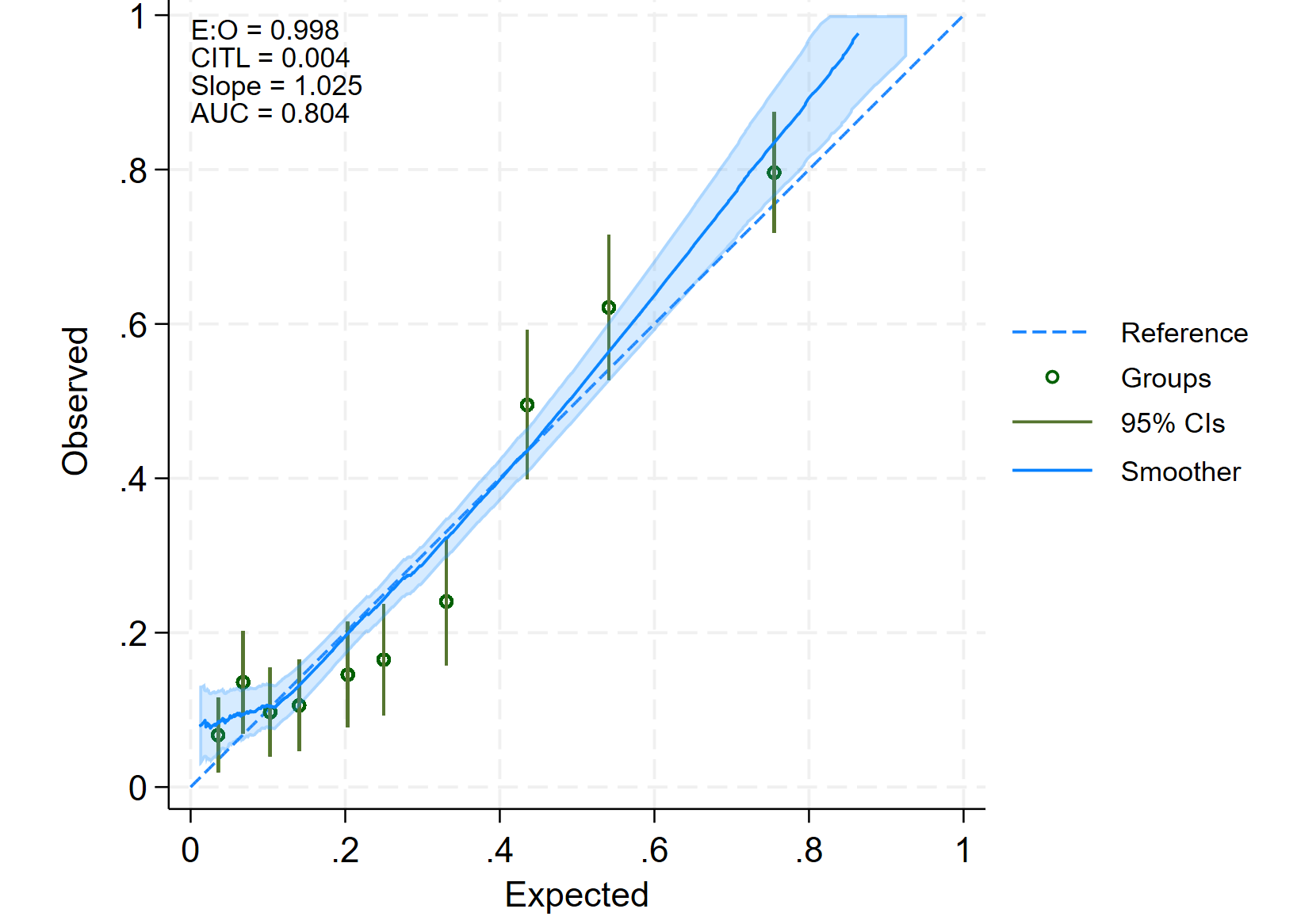

#### Figure S2. Calibration plot of Syrian anxiety symptom prediction model

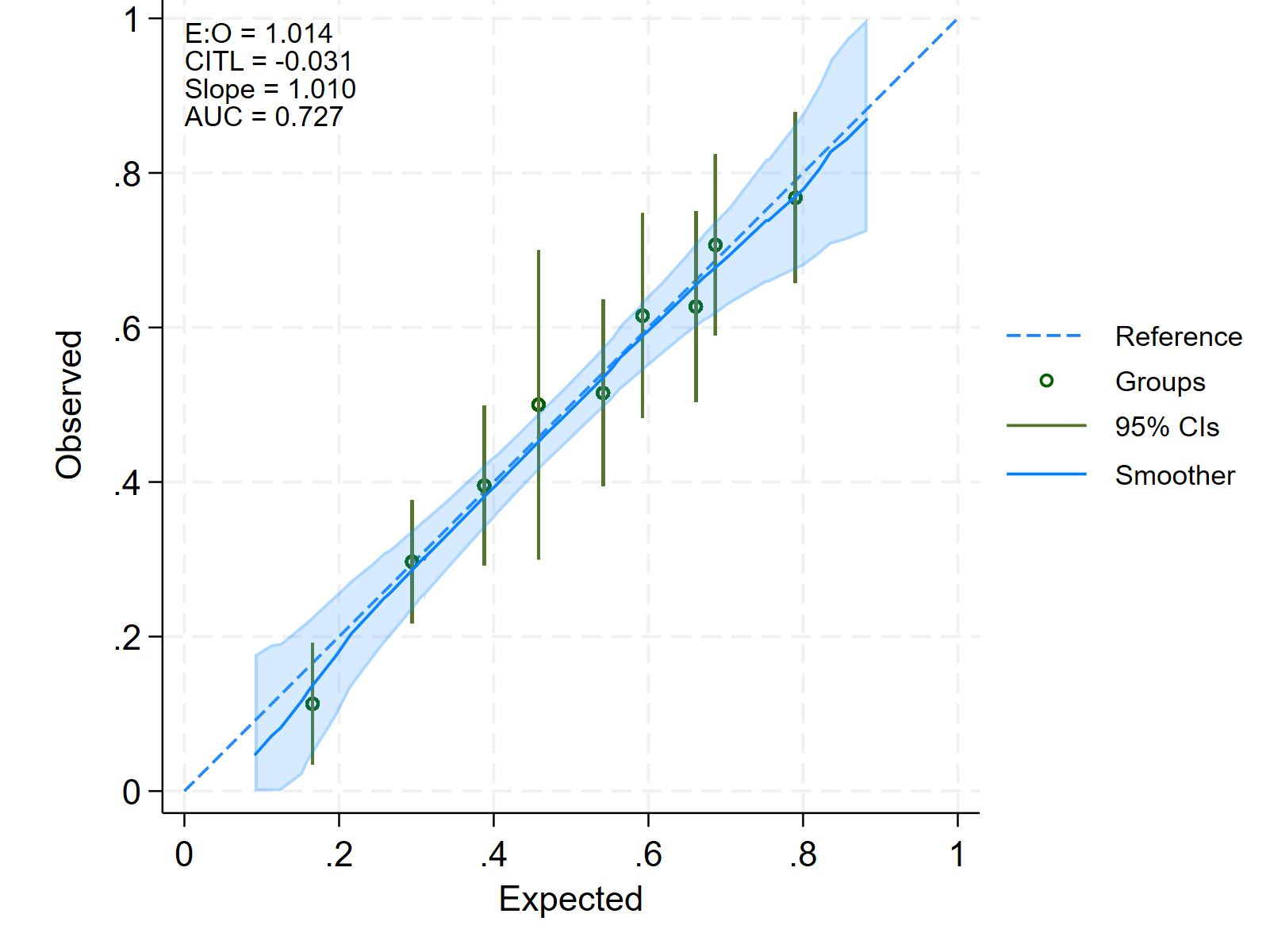
